# A proteomics workflow reveals predictive autoantigens in idiopathic pulmonary fibrosis

**DOI:** 10.1101/2021.02.17.21251826

**Authors:** Gabriela Leuschner, Christoph H. Mayr, Meshal Ansari, Benjamin Seeliger, Marion Frankenberger, Nikolaus Kneidinger, Rudolf A. Hatz, Anne Hilgendorff, Antje Prasse, Jürgen Behr, Matthias Mann, Herbert B. Schiller

## Abstract

**Rationale:** The diagnosis of idiopathic pulmonary fibrosis (IPF) requires exclusion of known underlying autoimmunity, as present in interstitial lung diseases associated with connective tissue diseases (CTD-ILD). However, autoantibodies of unknown significance have been repeatedly detected in IPF patients.

**Objectives:** We aimed to characterize autoreactivities in IPF patients beyond clinically established autoimmune panels by establishing an unbiased assay for de novo discovery of autoantigens in different forms of ILD and healthy controls.

**Methods:** We developed the proteomic Differential Antigen Capture (DAC) assay, capturing patient antibodies from plasma, followed by affinity purification of lung proteins coupled to mass spectrometry. Plasma antibodies from patients with IPF (n=35), CTD-ILD (n=24) and age-matched controls (n=32) were analyzed and validated in an independent cohort (IPF: n=40; CTD-ILD: n=20). Plasma antibody binding profiles were associated with clinical meta-data including diagnosis, lung function and transplant free survival.

**Measurements and Main Results:** We identified 586 putative autoantigens in both study cohorts with a broad heterogeneity among disease entities and cohorts. The prevalence of autoantibodies was higher in IPF compared to CTD-ILD. We identified a predictive autoimmune signature that was significantly associated with reduced transplant free survival in IPF. In particular, presence of autoantibodies to Thrombospondin 1 (THBS1) was associated with a significantly reduced survival in patients with IPF (p=0.002), independent of the study cohort, suggesting clinical relevance as predictive biomarker.

**Conclusions:** Unbiased proteomic profiling reveals that the overall prevalence of autoantibodies is similar in IPF and CTD-ILD patients and identifies novel IPF specific autoantigens associated with patient survival.

## Introduction

Idiopathic pulmonary fibrosis (IPF), the most common form of idiopathic interstitial pneumonia, is currently an incurable disease with a fatal prognosis of 2-5 years after diagnosis (Bjoraker et al., 1998). Although newly approved antifibrotic therapies (pirfenidone and nintedanib) can slow down disease progression, the only definitive therapy for IPF is lung transplantation (King et al., 2014; Noble et al., 2011; Richeldi et al., 2014). Autoimmunity-driven interstitial lung diseases, such as connective tissue disease related interstitial lung disease ILD (CTD-ILD), have a better prognosis and are characterized by the presence of auto-antibodies (e.g. Scl-70 antibody against topoisomerase-I in systemic sclerosis), which are used for diagnosis and classification of the patients. Diagnosis of IPF requires exclusion of all known causes for ILD including connective tissue diseases, which usually present with typical clinical signs and antibodies against various autoantigens. While absence of inflammation and autoimmunity is believed to be characteristic for IPF, there is a growing body of evidence that IPF may have clinical features compatible with an underlying autoimmune process. Circulating autoantibodies have been reported to be present in approximately one third of IPF patients (Lee et al., 2013; Leuschner et al., 2018; Moua et al., 2014), without meeting the required diagnostic criteria for CTD-ILD and interstitial pneumonia with autoimmune features (IPAF) (Fischer et al., 2015). Given the highly diverse clinical courses of IPF, comprehensive autoantibody profiles from IPF patients may benefit personalized therapy decisions.

It is even conceivable that the presence of autoantibodies and autoreactive T cells against unknown antigens could perpetuate pathology in IPF. Compared to healthy controls and patients with chronic obstructive pulmonary disease (COPD), patients with IPF show higher plasma levels of the B lymphocyte stimulating factor, which is essential for B-cell differentiation and B-cell survival as well as more C-X-C motif chemokine 13, which is needed for a regular B-cell migration in inflammatory tissue (Vuga et al., 2014; Xue et al., 2013). Furthermore, we previously identified an unexpected high prevalence of marginal zone B-cell-1 protein (MZB1) positive antibody-secreting plasma B-cells in ILD tissues, including IPF (Schiller et al., 2017). In our study, MZB1 levels correlated positively with tissue IgG and negatively with lung function parameters, suggesting a common association of IPF progression with antibody mediated autoimmunity (Schiller et al., 2017). Indeed, it has been shown that high levels of autoantibodies against vimentin (Li et al., 2017), heat-shock protein 70 (Kahloon et al., 2013), periplakin as well as anti-parietal cell antibodies (Beltramo et al., 2018; Taillé et al., 2011), were associated with a more severe disease in IPF.

In this study we aimed at the identification of disease specific and prognosis-relevant autoantibodies in IPF and CTD-ILD. We developed a proteomic workflow for high sensitivity and accuracy detection of lung disease associated autoantigens and compared antibody autoreactivities in independent CTD-ILD and IPF patient cohorts.

## Results

### A mass spectrometry workflow for proteome wide discovery of autoantigens in ILD

Recent advances approach autoantigen discovery in a large scale and high throughput manner using protein microarrays (Rosenberg and Utz, 2015), or phage display libraries, representing a synthetic human peptidome (Larman et al., 2011). These assays do however depend on synthetic antigens. To identify lung disease associated autoantigens derived from in vivo human tissue samples using unbiased mass spectrometry we developed the Differential Antigen Capture assay (DAC; see methods for detailed description), which is based on immunoprecipitation of human lung proteome extracts with patient immunoglobulins followed by quantitative shotgun proteomics similar to affinity purification (AP)-MS (Figure 1A). To characterize the sensitivity and specificity of our assay we analyzed plasma from patients with CTD-ILD, some of which were diagnosed with systemic sclerosis and tested positive by ELISA in clinical routine for Scl-70 auto-antibodies (specific antigen topoisomerase I; TOP1) (Figure 1B).

**Figure 1.**
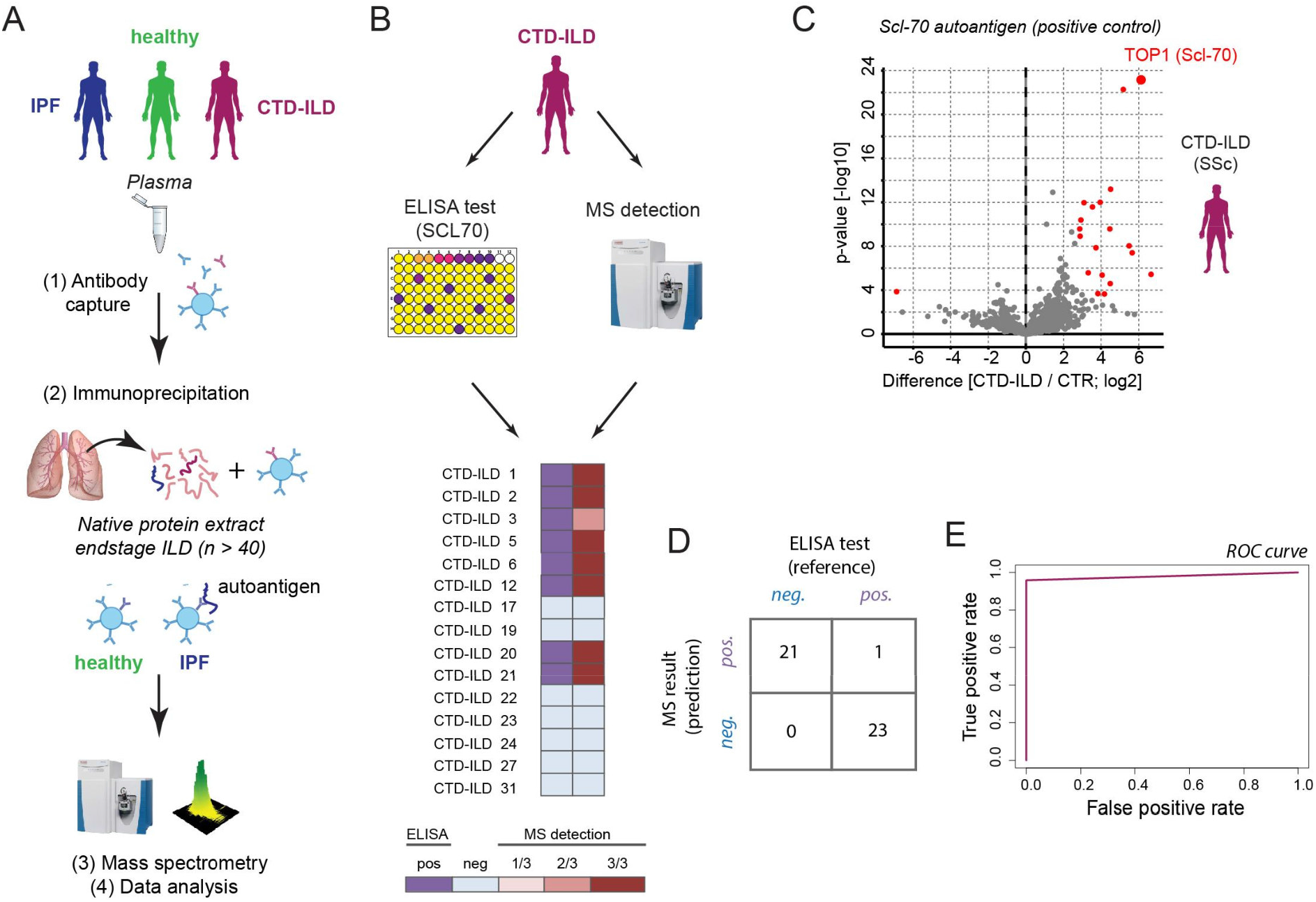
A proteomic workflow detects autoantigens with high sensitivity and specificity. **(A)** Experimental workflow. Antibodies, captured from plasma with protein-G beads, are used to precipitate proteins from native lung extracts of end-stage ILD patients. Differential protein binding to beads (disease versus healthy age matched controls) is quantified using state of the art shotgun MS-based proteomics. **(B)** Detection of plasma antibody reactivity to an autoantigen (SLC-70) by mass spectrometry was compared to a clinically used ELISA test. Some patients with connective-tissue related interstitial lung disease (CTD-ILD), tested positive for Scl-70 autoantibodies in the ELISA test. The color code indicates how many of the three replicates in MS-analysis showed a significant enrichment for Scl-70. **(C)** Representative volcano plot of a Systemic sclerosis (SSc) patient positive for the Scl-70 antigen (Toposisomerase 1; TOP1), shows enrichment of the autoantigen versus age matched healthy donors. Red dots indicate significantly enriched proteins (FDR <5%). **(D, E)** Each patient sample in panel B was measured in triplicates resulting in the depicted number of true and false positives. The specificity of significant Top1 (Scl-70) enrichment in ILD compared to age matched healthy donor controls by MS-analysis was 100%, while sensitivity was 96% (ROC analysis).

Our assay scores the proteins from native lung extracts as putative autoantigens in patients when they are significantly enriched (FDR<5%) on beads coupled to plasma antibodies from ILD patients versus the beads coupled to plasma antibodies from age matched healthy controls (Figure 1C). In comparison to patients who had negative results for Scl-70 antibodies (n=7), our assay identified clinically verified Scl-70-positive patients (n=8) with exceptionally good specificity and sensitivity, and equally good performance as the clinically approved ELISA test (Figure 1D,E). Of note, in contrast to the ELISA test our method simultaneously identified several other enriched putative autoantigens in these patients (Figure 1C). Thus, in this study we aimed to use the multiplexed and unbiased nature of the DAC assay to discover novel autoantigens in ILD and associate patient specific signatures with clinical features.

### Unexpected high prevalence of autoantibodies in IPF

To systematically compare the potential auto-reactivities of plasma antibodies in CTD-ILD and IPF, we performed the DAC assay in two independent study cohorts. Schematic of cohorts and availability of clinical data are shown in Figure 2.

**Figure 2.**
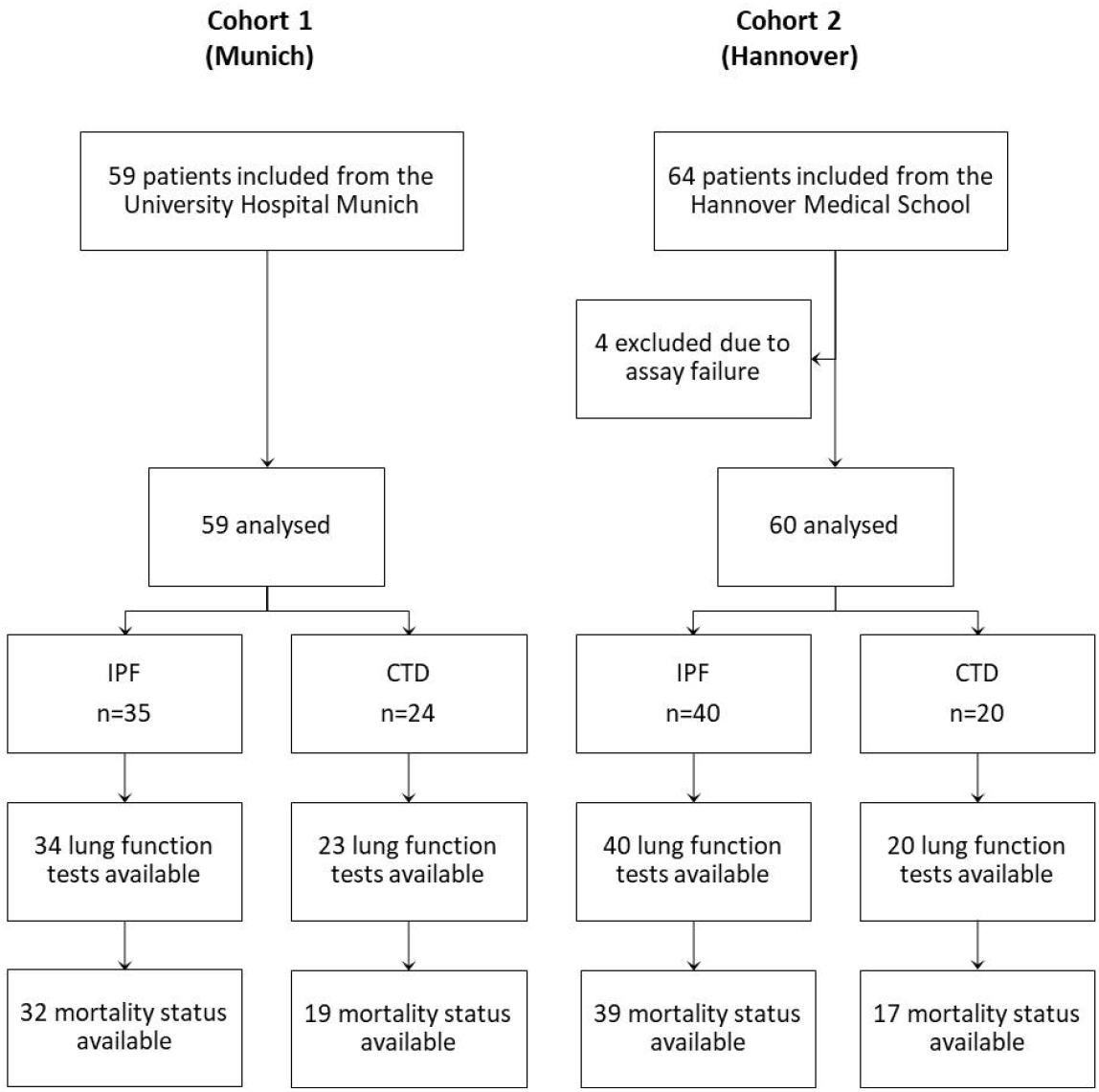
Two independent study cohorts used for autoantibody profiling from blood plasma.

Cohort-1 (Munich; Table 1) included IPF (n=35), CTD-ILD (n=24) and age-matched healthy controls (n=32; mean±SD 58.8±11.1 years, range 40-82 years). The CTD-ILD patients were diagnosed with systemic scleroderma (n=15), unspecific CTD (n=5), rheumatoid arthritis (n=4), and mixed connective tissue disease (n=1). The mean±SD age was 63.3±8.4 years in IPF and 62.7±10.2 years in CTD-ILD. Patients with IPF had significantly lower FVC % pred. than patients with CTD-ILD (54.5±15.8 vs. 65.7±18.9; p=0.025).

**Table 1:**
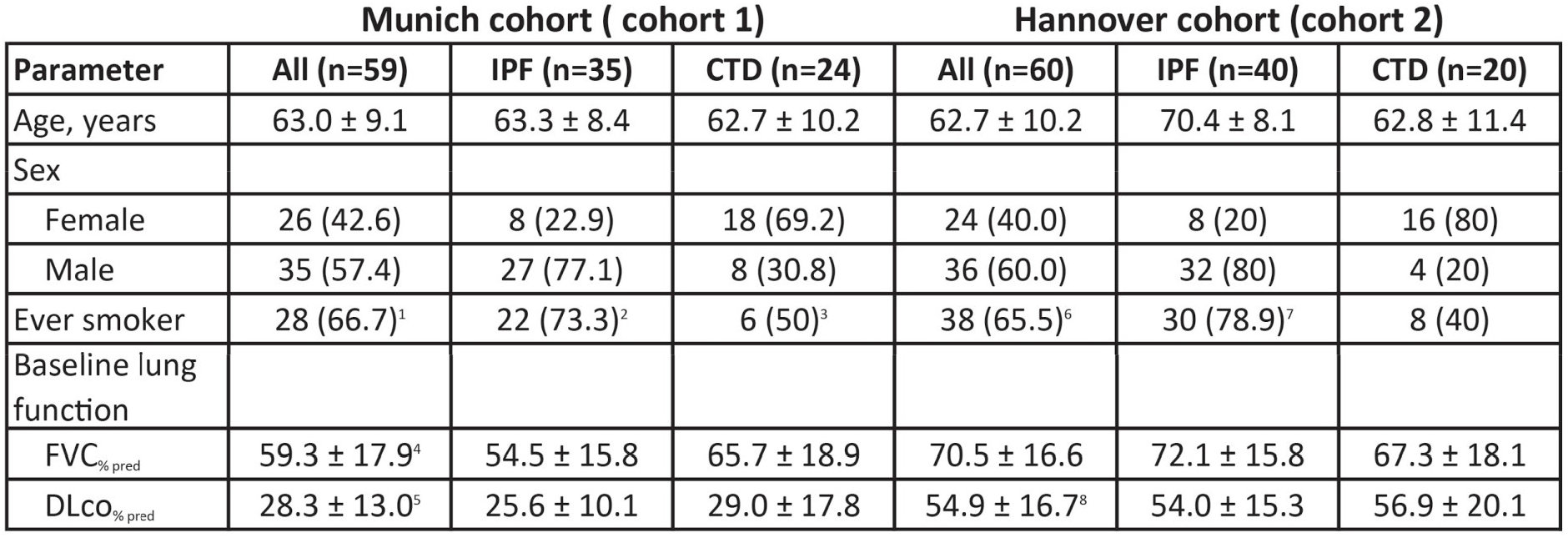
Baseline characteristics of patients included in the Munich and Hannover cohorts (cohort 1 and cohort 2). Data are presented as mean (±SD) or n (%). Abbreviations: percentage of forced vital capacity (FVC_%pred_), percentage of diffusion capacity of carbon monoxide (DLco_%pred_), ^1^ smoking status available in n=42, ^2^smoking status available in n=30, ^3^ smoking status available in n=12, ^4^ FVC available in n=59 (IPF n=34, CTD-ILD n=25), ^5^ DLCO available in n=49 (IPF n=28, CTD-ILD n=21), ^6^ smoking status available in n=58, ^7^ smoking status available in n=38, ^8^ DLCO available in n=54 (IPF n=38, CTD-ILD n=16).

| Munich cohort ( cohort 1) |  |  |  | Hannover cohort (cohort 2) |  |  |
| --- | --- | --- | --- | --- | --- | --- |
| Parameter | All (n=59) | IPF (n=35) | CTD (n=24) | All (n=60) | IPF (n=40) | CTD (n=20) |
| Age, years | 63.0 ± 9.1 | 63.3 ± 8.4 | 62.7 ± 10.2 | 62.7 ± 10.2 | 70.4 ± 8.1 | 62.8 ± 11.4 |
| Sex |  |  |  |  |  |  |
| Female | 26 (42.6) | 8 (22.9) | 18 (69.2) | 24 (40.0) | 8 (20) | 16 (80) |
| Male | 35 (57.4) | 27 (77.1) | 8 (30.8) | 36 (60.0) | 32 (80) | 4 (20) |
| Ever smoker | 28 (66.7)<sup>1</sup> | 22 (73.3)<sup>2</sup> | 6 (50)<sup>3</sup> | 38 (65.5)<sup>6</sup> | 30 (78.9)<sup>7</sup> | 8 (40) |
| Baseline lung function |  |  |  |  |  |  |
| FVC<sub>% pred</sub> | 59.3 ± 17.9<sup>4</sup> | 54.5 ± 15.8 | 65.7 ± 18.9 | 70.5 ± 16.6 | 72.1 ± 15.8 | 67.3 ± 18.1 |
| DLco<sub>% pred</sub> | 28.3 ± 13.0<sup>5</sup> | 25.6 ± 10.1 | 29.0 ± 17.8 | 54.9 ± 16.7<sup>8</sup> | 54.0 ± 15.3 | 56.9 ± 20.1 |

The second cohort (Hannover; Table 1) also included IPF (n=41) and CTD-ILD patients (n=20; sjogren’s syndrome n=6; systemic sclerosis n=5; rheumatoid arthritis n=5; CREST syndrome n=3; antisynthetase syndrome n=1). On average, with a mean±SD age of 70.4±8.1 years, IPF patients from cohort 2 were significantly older than all other patients (p=0.003) and patients from Hannover had a significantly better lung function than patients from Munich.

Using the DAC assay we quantified on average > 500 proteins per patient sample in all patient groups in both study cohorts (Figure S1A, see Table S1 and Table S2 for protein quantification of cohorts 1 (Munich) and cohort 2 (Hannover)). We found that putative autoantigens enriched over healthy age matched controls were mostly cytoplasmic, nuclear and cytoskeletal components (Figure S1B-D). However, we also identify several receptors and secreted proteins as putative autoantigens in ILD (Figure S1E). Interestingly, the distribution of these terms was similar between IPF and CTD-ILD. Adjusting for only significant proteins (significantly enriched in comparison to healthy controls), in cohort 1, we identified a mean±SD of 29±56 antigens (range 0 to 279) per patient in IPF and a mean of 12±19 (range 0 to 101) in CTD-ILD. With 6±5 antigens per patient in IPF and CTD-ILD, respectively, the average number of identified autoantigens in cohort 2 was distinctly lower. Taking together both cohorts, the mean±SD number of autoantigens per patient was 16±40 in IPF and 9±15 in CTD-ILD, revealing that surprisingly the prevalence of autoantibodies was not lower in IPF compared to autoimmune associated ILD (Figure 3A). Independent of the cohort, we recognized a broad heterogeneity of autoantigens in IPF: whereas the majority of identified autoreactivities in both IPF cohorts were only found in 1-2 patients, less than 10% of these antigens were enriched in 5 or more patients (Figure 3B).

**Figure 3.**
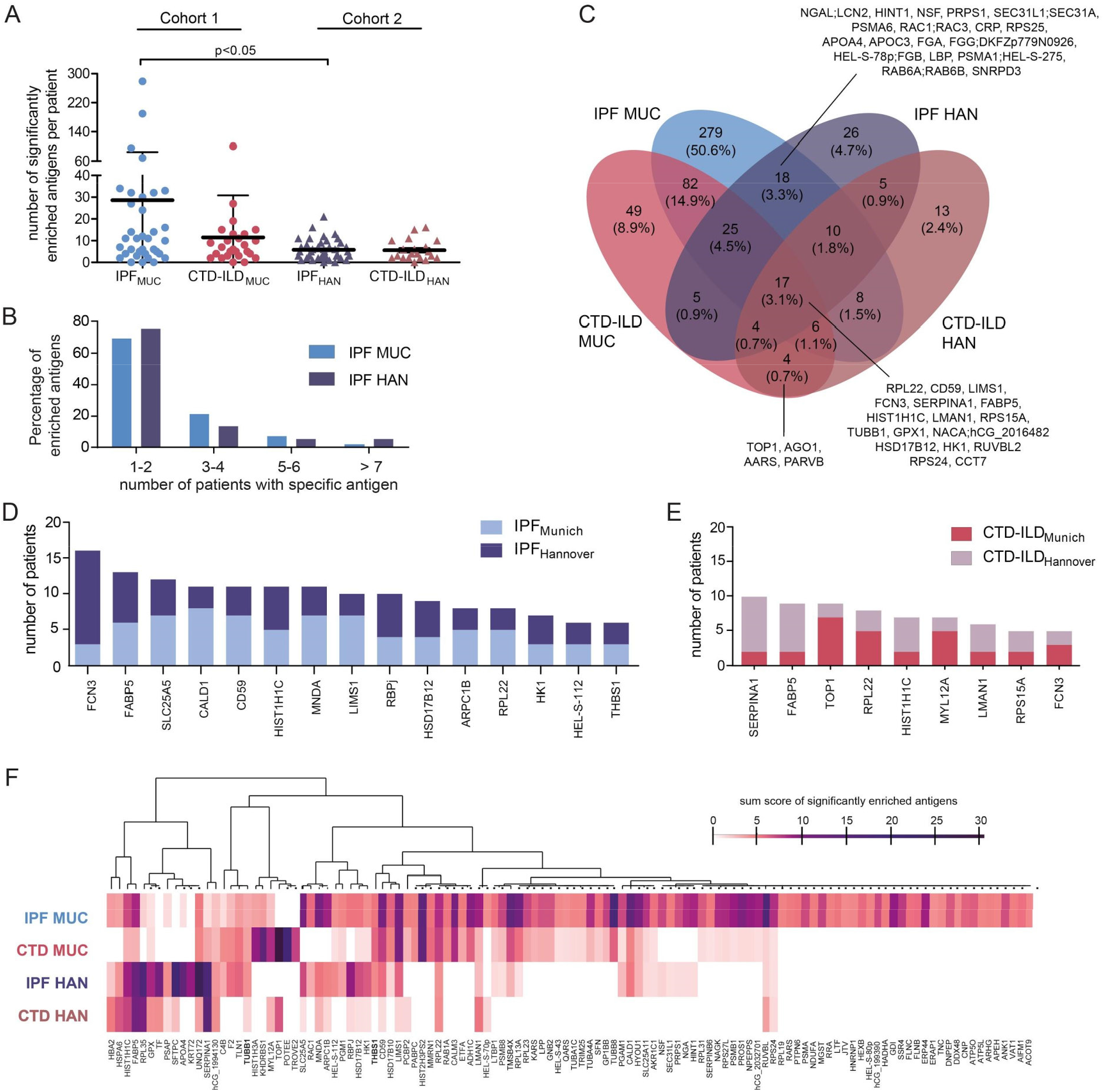
Common and individual autoantigens in IPF and CTD-ILD. **(A)** Number of significantly enriched autoantigens per patient in cohort 1 and cohort 2. **(B)** Percentage distribution of identified autoantigens in IPF cohort 1 and IPF cohort 2. **(C)** Venn diagram of individual and shared autoantigens in IPF and CTD-ILD across two cohorts (see Table S3 for Venn Diagram loadings). **(D)** Bar graph of the 15 most commonly detected autoantigens in IPF (present in at least three patients from each IPF cohort). **(E)** Bar graph of the nine most commonly found autoantigens in CTD-ILD (present in at least two patients from each CTD-ILD cohort). **(F)** Heat map of all four groups lists the top identified autoantigens that were significantly enriched in at least three patients in at least one group. Color code shows the score that is the sum of the Student’s T-test statistics of significant proteins (FDR <10%).

In IPF patients from cohort 1, the most commonly found autoantigens were RuvB-like 2 (RUVBL2; 28.6%; n=10), Histone H3 (HIST2H3PS2; 25.7%; n=9) and Tubulin beta-8 chain (TUBB8; 25.7%; n=9). RUVBL2 has previously been identified as a novel specific marker for systemic sclerosis, which was associated with male gender and older age (Kaji et al., 2014). Recently, it was also detected in anti-nuclear antibody negative systemic sclerosis (Pauling et al., 2018). So far, there is no data on RUVBL2 in IPF. The patients we tested positive for RUVBL2 autoantibodies here did not show any clinical signs for systemic sclerosis or CTD-ILD and were mainly male (70%). While autoreactivity to RUVBL2 and HIST2H3PS2 was not associated with any differences of FVC in IPF patients from cohort 1 (Figure S2A,B), patients with reactivity towards RUVBL2 showed a trend for better survival (p=0.052; Figure S2D). The detection of TUBB8 autoantibodies was associated with significantly higher FVC and a significantly better transplant-free survival in IPF patients from cohort 1 (Figure S2C,E).

In cohort 2, RUVBL2 and TUBB8 were not frequently detected as autoantigens in IPF. In contrast, the most commonly detected autoantigens were Ficolin-3 (FCN3; 32.5%; n=13), 60S ribosomal protein L35 (RPL35; 25%; n=10) and Alpha-1-antitrypsin (SERPINA1; 22.5%; n=9). None of these three antigens was associated with altered FVC levels (Figure S2F-H). Ficolin 3 is a recognition molecule in the lectin pathway of the complement system and expressed in the lung and the liver (Endo et al., 2007)(Munthe-Fog et al., 2009). In acute pulmonary inflammation, caused by LPS, alveolar FCN3 is increased (Plovsing et al., 2016).

Comparing significantly enriched autoantigens across diagnosis and patient cohorts showed substantial variation with most identified autoantigens specific to either a particular diagnosis or cohort (Figure 3C, see Table S3 for Venn Diagram loadings). We did however find 18 (3.3%) autoantigens, which were detected in both IPF cohorts and never in CTD-ILD. Specific for CTD-ILD in both cohorts we identified only four (0.7%) autoantigens, including TOP1. Combining both IPF cohorts (n=75), the distribution of the 18 IPF-specific autoantigens ranged from 2 (2.7%) to 9 patients (12.0%; Figure S3D, S3G-O). The identification of the Apolipoprotein A-IV (APOA4; n=9) was associated with significantly higher FVC values in comparison to the other IPF patients (p=0.016; Figure S3E). We are not aware of any data connecting APOA4 and lung fibrosis so far. A total of 17 autoantigens were shared in all 4 groups (Figure 3C and Figure S1F), and 15 autoantigens were present in at least three patients in each IPF cohort, with FCN3 being the most commonly detected (Figure 3D). We further identified 9 autoantigens that were present in at least two patients in each CTD-ILD cohort, including the CTD-ILD specific TOP1 with SERPINA1 having the highest prevalence (Figure 3E). Summing up the enrichment scores of significantly enriched proteins (FDR<10%), which were identified in at least three patients in one of the four study groups (cut-off sum score value >4), we identified 116 top autoantigens, which showed common and distinct association with study groups (Figure 3F). Of note, IPF patients from cohort 1, which was the clinically most severely affected group, had the largest autoantibody repertoire (Figure 3F).

### Autoantigen signature is associated with transplant free survival in IPF

To assess the predictive value of the autoantigens discovered in this study we integrated our data with clinical meta-data. We considered autoantigens present in at least three IPF patients. In total, 58 autoantigens met these requirements. In 11 patients (cohort 1: n=8; cohort 2: n=3) none of these 58 autoantigens were present. We performed Kaplan-Meier survival analysis for these 58 autoantigens, considering two groups either positive or negative for the respective autoreactivity. The mean±SD follow-up in the study cohort was 1.7±1.1 years. While in cohort 1, four patients died and 17 underwent lung transplantation, in cohort 2, ten patients died and none underwent lung transplantation.

We observed autoreactivities that predicted improved and worsened clinical outcome for the patients. On the one hand, patients with autorecativities towards APOA4 or Glutathione peroxidase 1 (GPX1) had a significantly better transplant-free survival (Figure S3A-B). On the other hand, our analysis identified autoreactivities to Thrombospondin-1 (THBS1; n=6; 8.0%; p=0.002; Figure 4A), Tubulin beta-1 chain (TUBB1; n=5; 6.7%; p=0.019; Figure 4B) and CD5L (n=3; 4.0%; p=0.0015; Figure 4C) as being associated with significantly reduced transplant-free survival time. Using multiple regression analysis and controlling for the confounding age, FVC and gender, we found that both THBS1 and TUBB1 were significant predictors for transplant-free survival (THBS1 HR: 3.98; 95%-CI: 1.432-11.074; p=0.008 - Table 3; TUBB1, HR 3.469; 95%-CI: 1.150 - 10.464; p=0.027). Importantly, THBS1 independently predicted survival in both study cohorts (Figure S3C-D). THBS1 is a secreted matricellular glycoprotein involved in cell-to-cell communication and latent TGF-beta activation (Murphy-Ullrich and Suto, 2018), which has implications in fibrotic diseases. TUBB1 is a gene that encodes for the tubulin β-1 chain (Burley et al., 2018). Together with alpha tubulins, the beta tubulins are the major constituents of microtubules, which are in turn major components of the eukaryotic cytoskeleton. CD5L is a key protein in the control of immune homeostasis and inflammatory disease (Sanjurjo et al., 2015).

**Table 3:** Multiple regression analysis in IPF patients of both cohorts combined for transplant-free survival. Abbreviations: forced vital capacity (FVC), hazard ratio (HR), 95% confidence interval (CI).

| variable | HR | 95%-CI | p-value |
| --- | --- | --- | --- |
| FVC | 0.959 | 0.939 – 0.981 | **<0.001** |
| Age | 0.621 | 0.952 – 1.030 | 0.621 |
| Gender | 1.603 | 0.554 – 4.639 | 0.384 |
| THBS1 | 3.98 | 1.432-11.074 | **0.008** |

**Figure 4.**
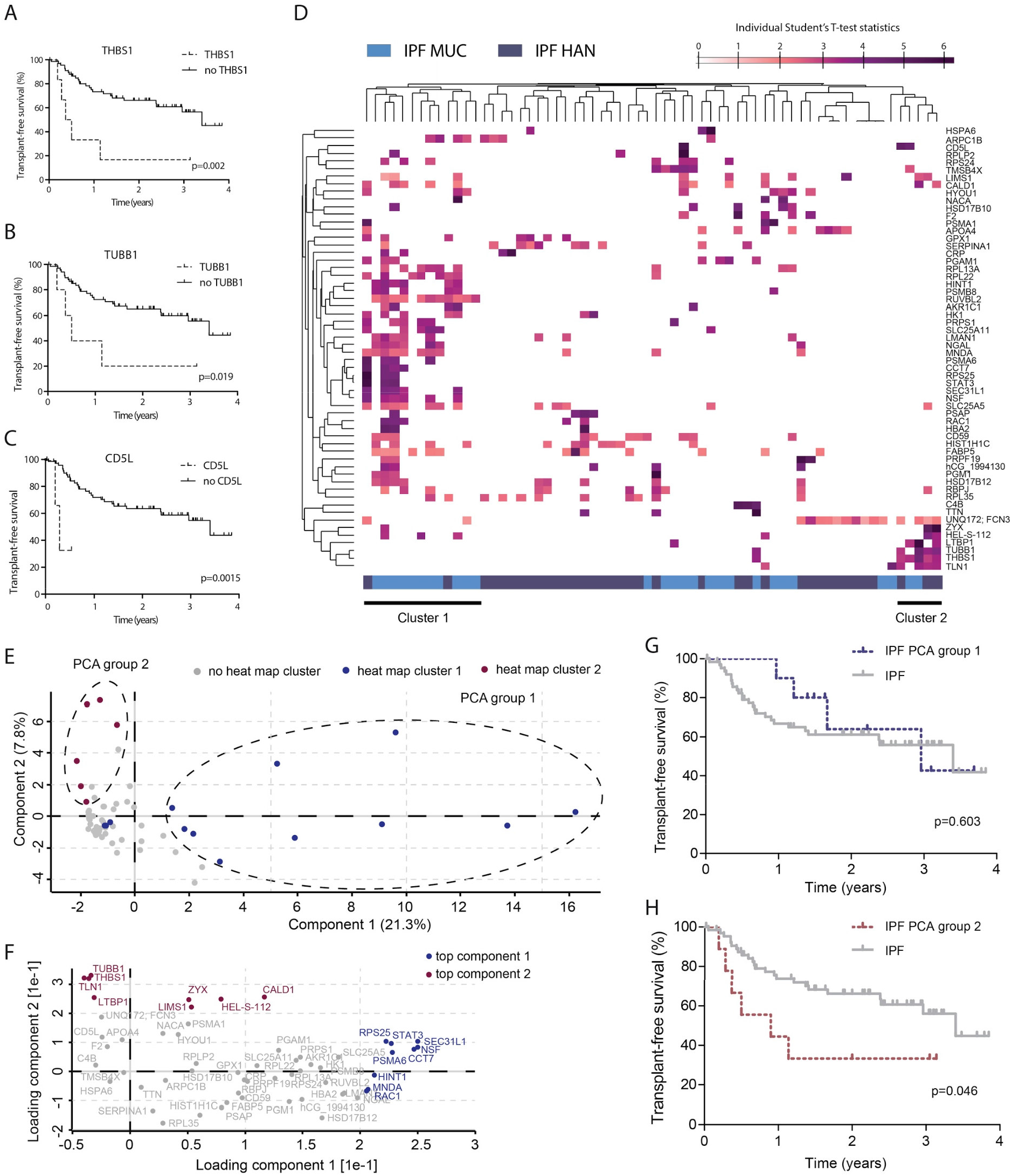
Identification of predictive autoantigens in IPF. **(A-C)** Kaplan-Meier analysis for transplant-free survival shows a significantly reduced survival in IPF patients with autoantibodies to THBS1, TUBB1, and CD5L respectively. **(D)** 58 proteins that were detected in both IPF cohorts and in at least two patients in one of the cohorts were grouped by hierarchical clustering. **(E)** Patients were grouped by principal component analysis with the same protein enrichment values used in (D). **(F)** Scatter plot showing the loadings of component 1 and component 2 of the PCA in (E). **(G, H)** Kaplan-Meier analysis for transplant-free survival does not identify differences in survival in IPF patients from patient group_1 in the PCA (G), but a significantly reduced survival in group_2 (H).

We observed that patients with autoreactivity for THBS1 often shared autoreactivity with TUBB1, which hinted towards a shared autoantigen repertoire in IPF patients at risk for mortality/lung transplantation. Displaying IPF patients from both cohorts in a heat map showed again the broad heterogeneity of autoantigens in both cohorts (Figure 4D). We detected two major clusters: while cluster 1 was based on 20 different autoantigens, cluster 2 was mainly driven by six autoantigens. These six genes were Talin 1 (TLN1), THBS1, TUBB1, Latent-transforming growth factor beta-binding protein 1 (LTBP1), Epididymis secretory protein Li 112 (HEL-S-112) and Zyxin (ZYX). TLN1 is a cytoplasmic protein essential for integrin activation (Lefort et al., 2012). LTBP1 is involved in the TGF-beta pathway (Miyazono et al., 1991).

Cluster 1 was formed by a group of 13 patients (cohort 1: n=11; cohort 2: n=2) and cluster 2 displayed six patients (cohort 1: n=3; cohort 2: n=3). Next, we used a principal component analysis (PCA) of the Z-scored Student’s T-Test statistics of the top 58 autoantigens to identify different groups of IPF patients. The analysis separated one group of patients in component 1 (group_1) and a second group in component 2 (group_2) (Figure 4E). All of the patients in group_1 (n=11) were from the heat map cluster 1 and six of nine patients (66.7%) from group_2 were from the heat map cluster 2. A scatter plot of the loadings of the PCA components revealed that component 1 was enriched for the genes Protein transport protein Sec31A (SEC31L1), Vesicle-fusing ATPase (NSF), T-complex protein 1 subunit eta (CCT7), Proteasome subunit alpha type-6 (PSMA6), Signal transducer and activator of transcription 3 (STAT3), 40S ribosomal protein S25 (RPS25), Histidine triad nucleotide-binding protein 1 (HINT1), Myeloid cell nuclear differentiation antigen (MNDA) and Ras-related protein Rac1 (RAC1), which were all but RAC1 present in the heat map cluster 1 (Figure 4F). Component 2 was mainly driven by TUBB1, THBS1, TLN1, LTBP1, ZYX, HEL-S-112, the LIM and senescent cell antigen-like-containing domain protein 1 (LIMS1) and Caldesmon (CALD1), with all but the last two autoantigens being present in heat map cluster 2.

Based on the PCA-groups, we again performed Kaplan-Meier survival curves for transplant-free survival. While patients from PCA-group 1 (cohort 1: n=8; cohort 2: n=3) did not show differences in comparison to all other IPF patients (Figure 4G), patients from PCA-group 2 (cohort 1: n=4; cohort 2: n=5) had a significantly shorter transplant-free survival in comparison to all other IPF patients from cohort 1 and 2 (median survival 0.9 years versus 3.4 years; p=0.046; Figure 4H). Of note, these patients did not differ significantly in terms of baseline FVC (p=0.155) and age (p=0.282) from the other IPF patients at time of analysis.

## Discussion

In this study we analyze antibody mediated autoimmunity in ILD. By developing the mass spectrometry based Differential Antigen Capture (DAC) assay we were able to globally compare the autoreactivities of plasma antibodies in CTD-ILD and IPF, which revealed that the IPF patients did surprisingly not have a lower prevalence of autoreactive antibodies. This analysis also enabled us to find novel autoantigens in IPF patients, some of which were significantly associated with transplant free survival in independent patient cohorts.

Many chronic lung diseases, including ILD, have been associated with deregulated type 2 immunity, which primarily evolved as defense against multicellular parasites (Gieseck et al., 2018). Type 2 immunity is essential for tissue repair processes and might be a common contributing factor in the development of fibrosis. The production of antibodies is a central feature of Type 2 immunity and it has been recognized that B-cell mediated immunity is not only important for host defense (e.g. against parasites) but may also contribute to tissue repair mechanisms (Allen and Sutherland, 2014). In the case of chronic insults to the organ, the permanent tissue damage and associated cytokine profiles may lead to a clonal evolution of autoreactive germinal centers that give rise to a self-amplifying autoimmune response, ultimately culminating in irreversible tissue fibrosis (Degn et al., 2017). This hypothesis has not been experimentally challenged in the context of ILD and it is completely unknown which clinical entities of ILD have a causal association with the production of autoantibodies. Our newly developed autoantigen discovery workflow is highly competitive with current state of the art workflows in several respects: (1) We only needed minute amounts of plasma samples (20-50 μl per patient). (2) A non-targeted approach was used. We fed the immunoprecipitation assay with a full representation of the diseased lung proteome and identified antigens using mass spectrometry. Targeted assays depend on recombinant purified proteins, making a comprehensive and high throughput screening difficult. (3) Autoantigens can be a consequence of disease associated posttranslational protein modifications (PTMs), which also cannot be easily addressed with methods based on recombinant proteins. We are therefore confident that the DAC assay offers a number of advantages for both research and future clinical utility. For instance, the DAC assay can be used to shed light on the currently ongoing discussion on a potential role of autoantibodies in (long) COVID-19 (Khamsi, 2021).

Our data shows that there is a great heterogeneity of autoantibody profiles in IPF and CTD-ILD. We detected autoreactivities that were diagnosis-specific, but also a large number of shared autoantigens across CTD-ILD and IPF. Autoantibodies against THBS1, which were found in 8% of IPF patients, were an independent predictor for transplant-free survival in IPF patients of both cohorts, when adjusting for baseline age, FVC and gender. THBS1 is a glycoprotein with multiple functions, including the inhibition and stimulation of angiogenesis and tumor progression or wound healing (Bornstein, 1995). It has been detected in several cell types like macrophages, fibroblasts, vascular smooth muscle cells, endothelial cells and platelets (Ide et al., 2008). As THBS1 seems to be an important activator of TGFb-1 (Crawford et al., 1998; Murphy-Ullrich and Poczatek, 2000), a major role of THBS1 in fibrotic disease is discussed. The plasma levels of THBS1 have been shown to be significantly elevated in patients with IPF in comparison to healthy controls and sarcoidosis patients (Ide et al., 2008), with an inverse correlation with FVC. We identified several other autoantigens with association with the THBS1/TUBB1 signature, including TLN-1, LTBP1, HEL-S-112, ZYX, LIMS1 and CALD1. Like THBS1, some of these proteins have also been recognized in terms of fibrotic disease.

We acknowledge the limitations of our study. First, we included two independent cohorts, which differed in several ways. IPF patients from cohort-2 were not only older but also more healthy compared to patients from cohort-1. Moreover, the body fluids we used from cohort 1 were plasma samples and the ones from cohort-2 serum, which might account for some of the heterogeneity between cohorts. The sensitivity of our mass spectrometry workflow depends on several parameters that can vary for the respective autoantigen/antibody pairs. The enrichment of autoantigen depends on (1) autoantibody titer in plasma, (2) antibody affinity, (3) concentration of autoantigen in the tissue extract, (4) other unknown criteria. Since we had to choose one defined condition for our experiments we expected to miss some of the possible targets. Indeed, we did not identify autoantigens that have previously been shown to be associated with disease severity or survival in IPF, such as anti-vimentin, anti heat-shock protein 70 or anti-parietal cell antibodies (Beltramo et al., 2018; Kahloon et al., 2013; Li et al., 2017; Taillé et al., 2011).

In conclusion, we have developed a novel proteomic assay for unbiased autoantibody profiling that can be used to discover autoimmunity in various clinical scenarios. The surprisingly high prevalence of autoantibodies in IPF warrants further research on the causal roles of these autoimmune reactions in disease progression. We identify a set of IPF patients with specific autoantibody profiles, most notably to the secreted glycoprotein THBS1, that show reduced survival in two independent study cohorts.

## Supporting information

Table S1_MS table cohort 1 (Munich)

Table S2_MS table cohort 2 (Hannover)

Table S3_Venn Diagram loadings

## Data Availability

Proteome raw data and MaxQuant processing tables can be downloaded from the PRIDE repository (Perez-Riverol et al., 2019) under the accession number PXD024113 (Munich, cohort 1) and PXD024123 (Hannover, cohort 2).

## Supplementary information

**Figure S1.**
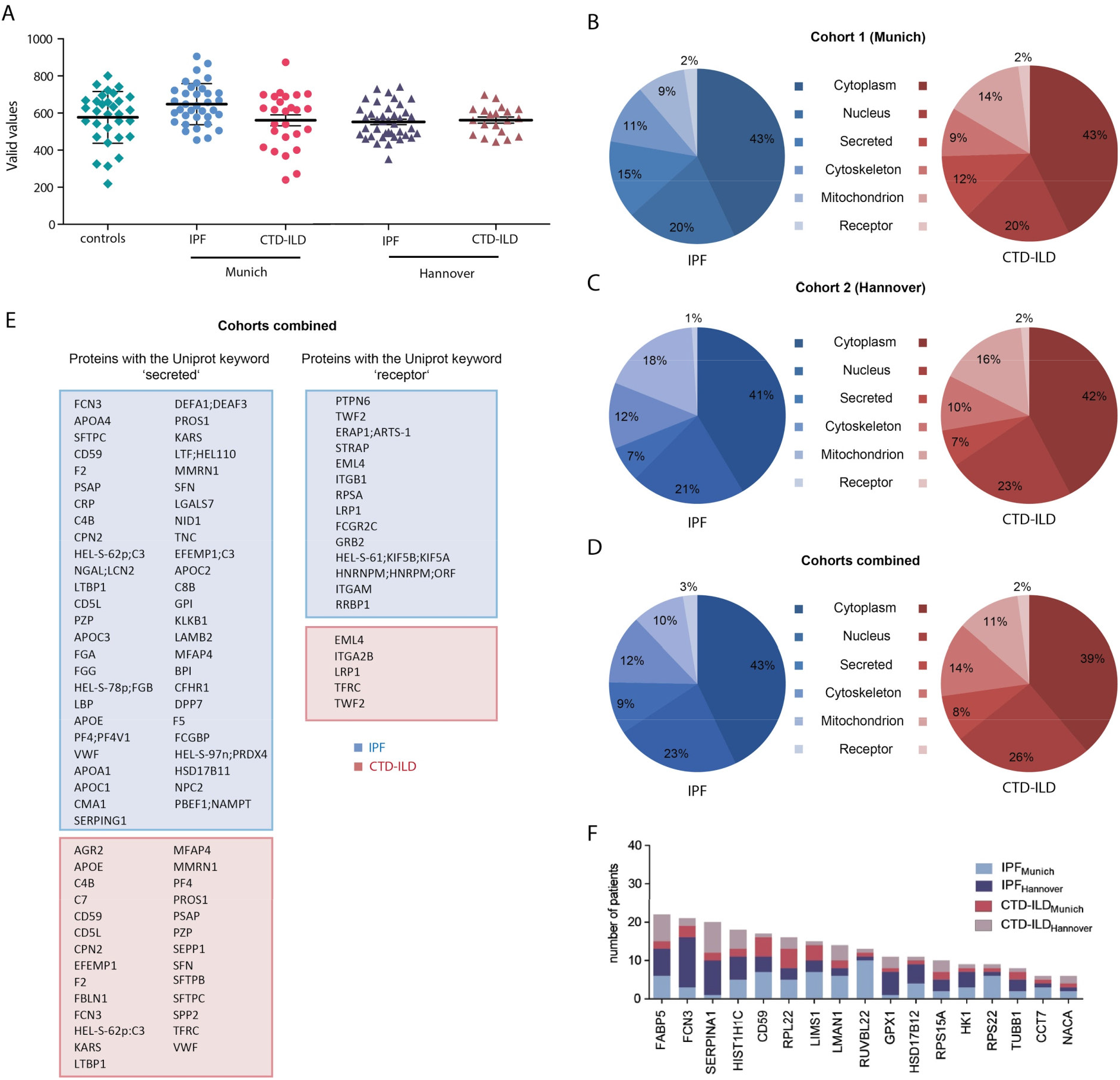
Protein quantification and GO terms of enriched autoantigens. **(A)** Number of quantified proteins across experimental groups and cohorts. **(B - D)** Categories of Autoantigens identified in IPF and CTD-ILD showed similar distribution, in cohort 1 (Munich) **(B)**, cohort 2 (Hannover) **(C)**, and when both cohorts combined **(D). (E)** Autoantigens in both cohorts combined, that correspond to the Uniprots keywords “secreted” and “receptor”; blue corresponds to IPF and red to CTL-ILD patients. **(F)** Bar graph showing the distribution of the 17 most common identified autoantigens, which were shared in all four groups, of IPF and CTD-ILD from both cohorts.

**Figure S2.**
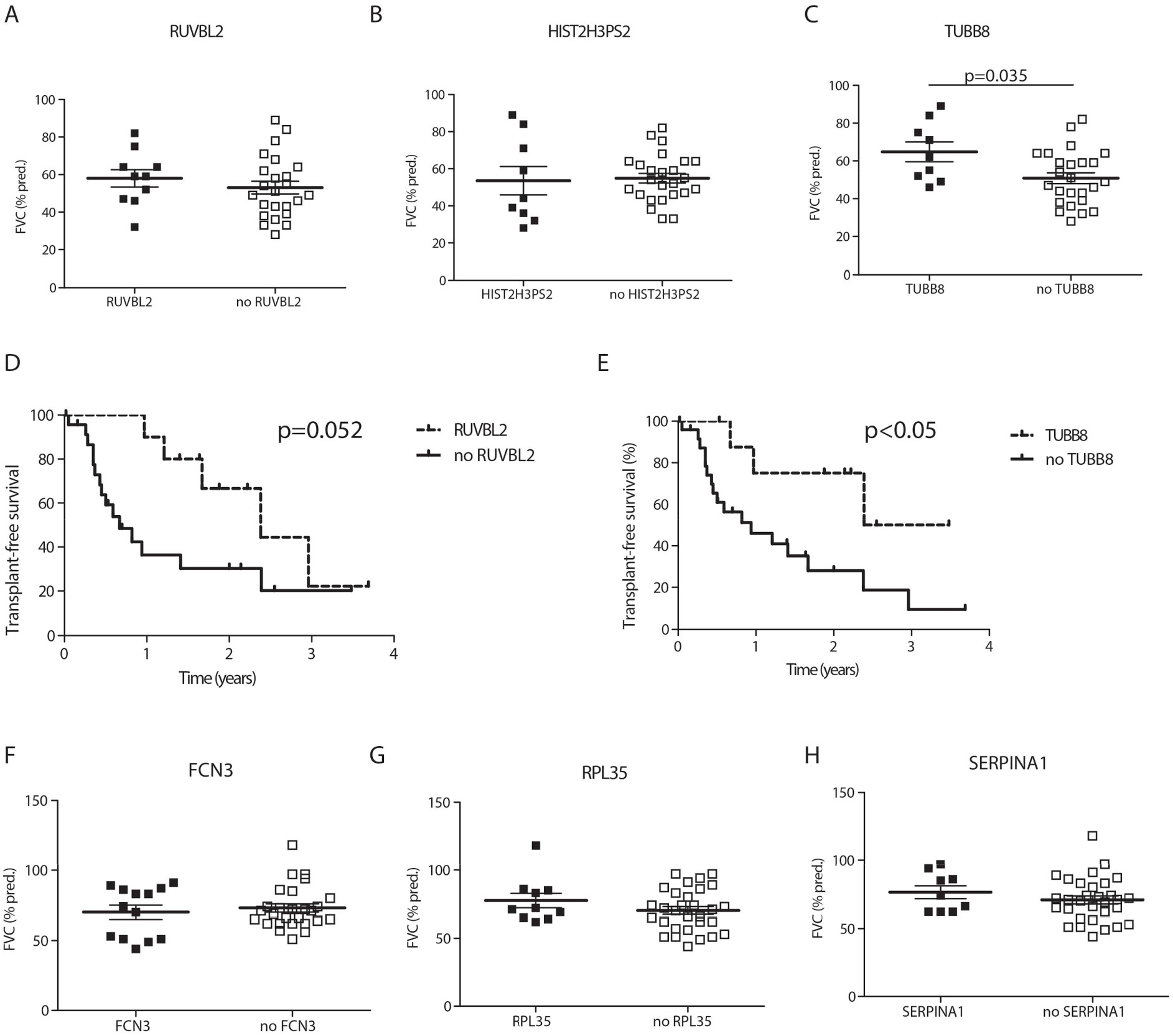
Most common identified autoantigens in IPF from cohort 1 and cohort 2. **(A - C)** RUVBL2, HIST2H3PS2 and TUBB8 were the most commonly found autoantigens in IPF. Only TUBB8 detected significant different FVC distribution. **(D)** Kaplan-Meier survival showed a tendency for better transplant-free survival in RUVBL2 positive patients. **(E)** Kaplan-Meier survival analysis identified better survival in TUBB8 positive patients. **(F - H)** FCN3, RPL35 and SERPINA1 were most commonly found in IPF from cohort 2 but showed no differences in FVC.

**Figure S3.**
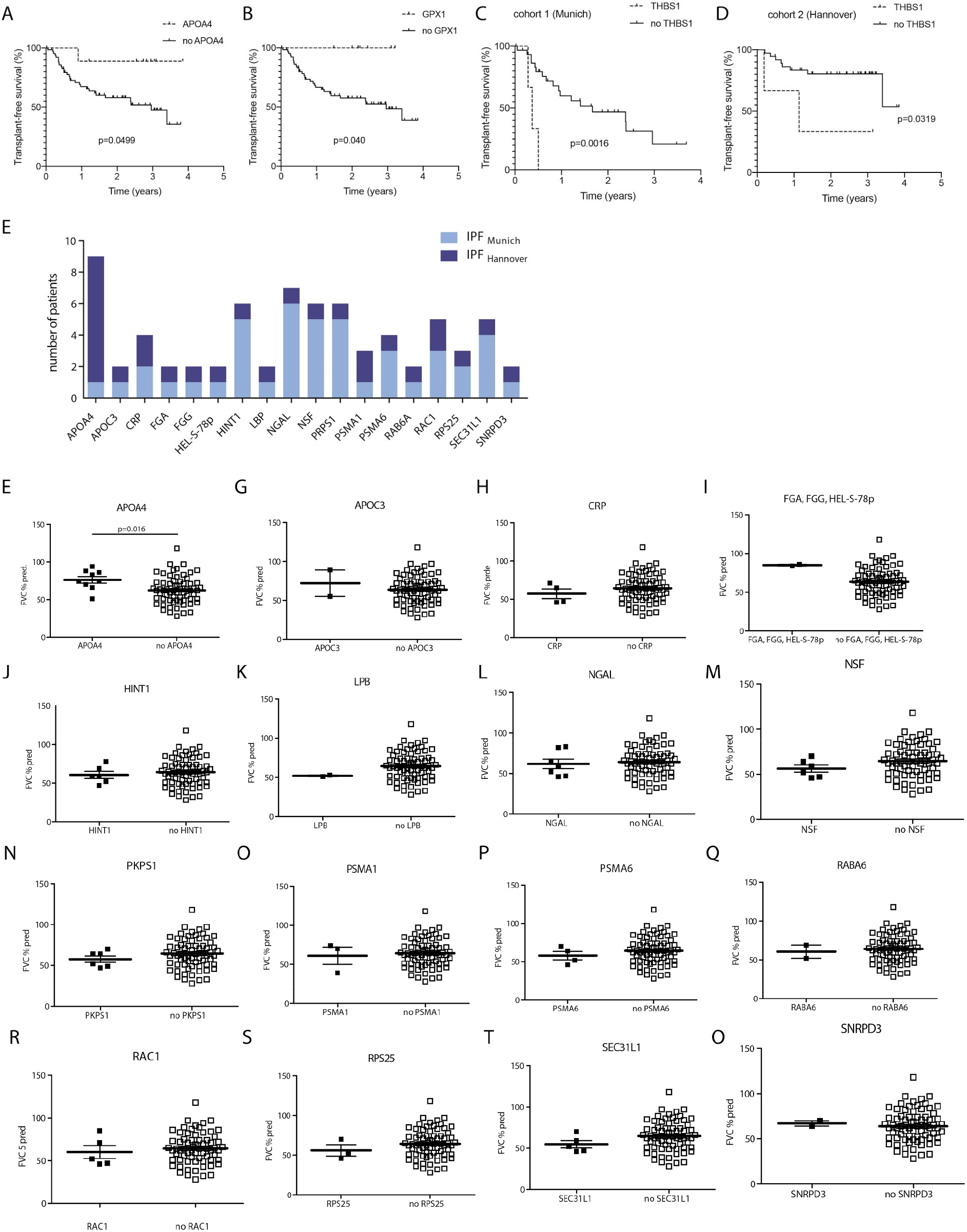
Identified IPF-specific autoantigens. **(A-B)**Kaplan-Meier survival curves for autoantibodies to APOA4 and GPX1 in IPF patients from both cohorts combined. **(C-D)** Kaplan-Meier survival curves show a significantly worse survival in IPF patients with autoantibodies against thrombospondin-1 (THBS1) in both cohort 1 (C), and in cohort 2 (D). **(E)** Distribution of 18 autoantigens which were identified in cohort 1 (light blue) and cohort 2 (dark blue). **(E-O)** FVC of patients with autoantigens in comparison to all other IPF patients identified APOA4 to be associated with significantly higher FVC.

## Supplementary Tables

**Table S1:**

Protein quantification results from the DAC assay for cohort 1 (Munich)

**Table S2:**

Protein quantification results from the DAC assay for cohort 2 (Hannover)

**Table S3:**

Protein loadings of the disease and cohort groups for the Venn Diagram in Figure 3C.

## Material and Methods

### Study cohort

Human lung tissues, derived from lung transplantation (donor and ILD recipients), were obtained from the BioArchive of the Comprehensive Pneumology Center Munich (CPC-M).

The autoantibody study population consisted of two independent ILD cohorts. Plasma samples of study cohort 1 (Munich cohort) were derived from IPF and CTD-ILD patients from the BioArchive of the CPC-M. Age-matched, healthy controls without signs of infection or respiratory symptoms were also received from the BioArchive of the CPC-M. Serum samples of the study cohort 2 (Hannover cohort) were obtained as part of a cooperation of the German Centers for Lung Research (DZL).

Blood samples were either collected in the respective ILD outpatient clinic during routine visits or in the inpatient unit during evaluation for lung transplantation. Diagnosis of IPF was made by high resolution computed tomography (HRCT), or, if available, histopathological findings from surgical lung biopsies (SLB) according to the official ATS/ERS/JRS/ALAT statement on the diagnosis and management of IPF (Raghu et al.; 2011). Patients with CTD-ILD had a radiographic proven ILD with an underlying systemic sclerosis, sjögren syndrome, rheumatoid arthritis, polyarthritis or undifferentiated CTD.

All patients/healthy controls had to be older than 18 years and all gave written informed consent. The study was performed according to the local ethics committee of the Ludwig-Maximilian University Munich (approval number 333-10 and 382-10). Patients gave written informed consent to the DZL broad-consent form and the study was approved by the local ethics committee of the Medizinische Hochschule Hannover (2923-2015).

### Human patient material processing

For plasma sampling, fresh venous blood was collected in EDTA-coated vacutainer tubes (Sarstedt, Nümbrecht, Germany). After centrifugation, supernatant plasma was separated from blood cells and immediately stored at −80°C degree in the CPC BioArchive. For cohort 2, Serum samples were obtained from the collaborators in Hannover, and stored at −80°C.

Protein extraction of end stage lung tissue was performed from explanted lungs of 41 ILD patients and 12 healthy donors undergoing lung transplantation. After lung transplantation, the lung tissue is collected in our CPC BioArchive in a standardized way. Lung tissue is cut into pieces of 0.5 cm^**3**^, snap frozen in liquid nitrogen and stored at −80°C degree. For protein extraction, frozen lung tissue pieces were subject to cryo-bead milling in TissueLyser2 (Eppendorf). The resulting powder was resuspended in RIPA-buffer (50mM Tris HCl pH 7.4, 150 mM NaCl, 1% Triton X100, 0.5% sodium deoxycholate, 1 mM EDTA, 0.1% SDS) supplemented with protease inhibitor and a phosphatase inhibitor cocktail (both Roche; Penzberg, Germany) and assisted by sonication (10 cycles, 30sec power, 30 sec pause) (Bioruptor, Diagenode). Samples were incubated on ice for 30 minutes and undissolved debris was removed by centrifugation for 5 minutes at 18000g. Bicinchoninacid Assay (BCA) (Pierce), was used to analyse protein concentration. For the protein extraction pool, equal amounts of all samples were pooled.

### Differential Antigen Capture assay (DAC)

To identify autoantigen-autoantibody interactions in ILD, we developed the Differential Antigen Capture assay (DAC), based on immunoprecipitation. Protein G agarose-coupled beads (Pierce, Thermo Fisher Scientific, 20399), which can bind up to 11-15mg human IgG per ml of settled resin (50% of slurry), according to manufacturer information, were used to catch antibodies from plasma. Protein extract from ILD and control patient lung tissue was added subsequently to capture antigens, binding to the antibodies on the beads. Immunoprecipitation was performed in triplicates for each patient.

Plasma was thawed on ice and centrifuged at 16.000xg for 5 minutes at 4°C degree. As described above, antigen protein extraction was derived from end stage lung tissue from explanted lungs of 41 ILD patients and 12 healthy donors undergoing lung transplantation, and equal amounts of all samples were pooled. Protein extractions were also thawed on ice.

First, 20µl of Protein G agarose beads were put in 96 well-filter plates (MultiScreenHTS-BV, 1.2 µm, Millipore) and washed with 200µl wash buffer (0.1% IPGAL, 5% Glycerol, 50 mM Tris pH 7.4, 150 mM NaCl, Roche EDTA free protease inhibitor) by centrifugation at 100g for 1 minute. Beads were then loaded with a mixture of 195µl of wash buffer and 5µl of plasma (MUC cohort)/serum (Hannover cohort) supernatants, and incubated for 1 hour at room temperature, at 900 rpm shaking. Afterwards, supernatant was removed from the filter plates by centrifugation at 100g for 1 minute and the beads were washed 3 times with 200µl wash buffer.

Antibody saturated beads were loaded with 150µg of antigen containing protein extract in 200µl wash buffer and incubated for 1 hour at room temperature shaking at 400 rpm. Filter plates were washed with wash buffer (without protease inhibitor) three times and then three times with PBS to prepare for proteomic digestion.

### Mass Spectrometry

Immunoprecipitation samples were subject to on-bead digest. Beads were incubated in digestion buffer (50 µl 8 M Urea Hepes pH8, 0.5µg LysC (Wako, #129-02541, 10 AU, in ABC), 10 mM DTT) for 1 hour at room temperature in a shaker (600 rpm). Afterwards, 0.5µg Trypsin (Trpysin from porcine pancreas: Sigma-Aldrich, T6567) was added in 200µl 50 mM ABC with 55 mM CAA and beads were again incubated for 1 hour at room temperature gently shaking (600rpm). 96 well filter plates were centrifuged at 100g for 1 minute and digested peptides as flow-through were collected in clean 96 well plates. Filters were washed again with 50µl quenching buffer [2M Urea, 50 mM Thiourea, 2mM Hepes in 50 mM ABC (= 50mM NH4HCO3)] and centrifuged at 100g for 1 minutes. Digestion continued overnight at 37°C and 600rpm and was stopped by acidifying the samples to 1% TFA.

Peptides were purified using stage-tips containing a poly(styrene-divinylbenzene) copolymer modified with sulfonic acid groups (SDB-RPS), as previously described (Schiller et al., 2015)(Kulak et al., 2014). Stage tips were first activated with 100µl ACN. For equilibration, we first ran 100µl 30%MeOH, 1% TFA over the stage tips and then 200µl 0.2% TFA. The samples were loaded in 1 % TFA. Afterwards, stage tips were first washed twice with 100µl isopropanol in 1% TFA and then again with 200µl 0.2% TFA. Finally, samples were eluted with 60µl of 5% Ammonia and 80% ACN. Afterwards, samples were evaporated at 30°C degree (Eppendorf Evaporater Plus). The final eluates were dissolved in 6 µl buffer A* (MS-loading buffer) under sonication.

Approximately 1 μg of peptides were separated in one hour gradients on a 50-cm long (75-μm inner diameter) column packed in-house with ReproSil-Pur C18-AQ 1.9 μm resin (Dr. Maisch GmbH). Reverse-phase chromatography was performed with an EASY-nLC 1000 ultra-high pressure system (Thermo Fisher Scientific), which was coupled to a Q-Exactive Mass Spectrometer (Thermo Scientific). Peptides were loaded with buffer A (0.1% (v/v) formic acid) and eluted with a nonlinear 240-min gradient of 5–60% buffer B (0.1% (v/v) formic acid, 80% (v/v) acetonitrile) at a flow rate of 250 nl/min. After each gradient, the column was washed with 95% buffer B and re-equilibrated with buffer A. Column temperature was kept at 50 °C by an in-house designed oven with a Peltier element (Thakur et al., 2011) and operational parameters were monitored in real time by the SprayQC software (Scheltema & Mann, 2012). MS data were acquired with a shotgun proteomics method, where in each cycle a full scan, providing an overview of the full complement of isotope patterns visible at that particular time point, is follow by up to ten data-dependent MS/MS scans on the most abundant not yet sequenced isotopes (top10 method) (Michalski et al, 2011a). Target value for the full scan MS spectra was 3 × 10^6^ charges in the 300−1,650 *m/z* range with a maximum injection time of 20 ms and a resolution of 70,000 at *m/z* 400. The resulting mass spectra were processed using the MaxQuant software (Cox and Mann, 2008), which enabled label free protein quantification (Tyanova et al., 2016). As previously described (Schiller et al., 2015), peak lists were searched against the human Uniprot FASTA database (*November 2016*), and a common contaminants database (247 entries) by the Andromeda search engine (Cox et al., 2011).

### Clinical data

Clinical parameters were obtained at the time when the plasma was collected and included demographics (age, gender, smoking status, lung function [forced vital capacity (FVC) (% pred.), FVC (l), and diffusing capacity of the lung for carbon monoxide (DLCO) (SB) (% pred.)]. Transplant-free survival was retrospectively evaluated after plasma sampling.

### Data analysis

Statistical analysis of clinical data included t-test statistics, ANOVA tests, Fisher’s exact test and Kaplan-Meier survival analysis of lung transplantation-free survival using the GraphPad Prism 5 software.

To identify the most commonly and robustly found autoantigens, we used a score based on the sum of the Student’s T-test statistics values: ILD patients were tested against control patients, and of the significantly enriched proteins (FDR<10%), the score was calculated, per patient per protein (sum score of significantly enriched antigens). Antigens were only included if they were identified in at least three patients in at least one of the four study groups and a cut-off sum score value >4 was used.

All other statistical and bioinformatics operations (such as normalization, data integration, annotation enrichment analysis, hierarchical clustering, principal component analysis), were run with the Perseus software package (version 1.5.3.0 and 1.6.2.3. and 1.6.10.50) (Tyanova et al., 2016).

## Author contributions

HBS conceived the research narrative and supervised the entire study. HBS and GL wrote the paper. CHM and GL performed proteomics experiments. GL analyzed clinical data. GL, CHM, and MA analyzed proteomics data. BS, NK, MF and RAH collected clinical samples. AH, AP, JB, MM and HBS provided resources. All authors read and approved the manuscript.

## Acknowledgements

We thank all the patients and their families for supporting the progress of science. We gratefully acknowledge the provision of human biomaterial and clinical data from the CPC-M bioArchive and its partners at the Asklepios Biobank Gauting, the Klinikum der Universität München and the Ludwig-Maximilians-Universität München. We thank Ina Koch, Britta Peschel and Annika Frank for managing the Asklepios Biobank and the CPC-M Bioarchive and supporting this study. We thank Igor Paron and Christian Deiml for expert support of the proteomics pipeline. This work was supported by the German Center for Lung Research (DZL), the Helmholtz Association and the Max Planck Society.

